# Recruitment and Retention in mhealth Interventions for Addiction and Problematic Substance Use: A Systematic Review

**DOI:** 10.1101/2023.09.09.23295094

**Authors:** Bruce Kidd, Jessica C McCormack, David Newcombe, Katie Garner, Azim O’Shea, Gayl Humphrey

**Author notes:** **Address Correspondence** Gayl Humphrey, National Institute for Health Innovation, School of Population Health, University of Auckland, Private Bag 92019, Auckland 1142, Aotearoa | New Zealand.

## Abstract

**Background:** Disordered and problematic addictions are significant public health issues. It has been proposed that mHealth interventions can provide new models and intervention delivery modalities. However, research shows that studies that evaluate mHealth interventions for addiction disorders have low recruitment and high attrition. This study aims to identify published peer-reviewed literature on the recruitment and retention of participants in studies of mHealth interventions for people with addiction or problematic use and to identify successful recruitment and retention strategies.

**Methods:** Relevant studies were identified through Medline, Embase, PsychINFO, and Cochrane Central Register of Controlled Trials (CENTRAL) after January 1998. Studies were limited to peer-reviewed literature and English language published up to 2023. The revised Cochrane Risk of Bias RoB 2 tool was used to assess the risk of bias.

**Results:** Of the 2135 articles found, 60 met the inclusion criteria and were included. The majority of studies were for smoking cessation. Only three studies retained 95% of participants at the longest follow-up, with ten studies retaining less than 80% at the longest follow-up, indicating a high risk of retention bias. Those studies with high retention rates used a variety of recruitment modalities; however, they also recruited from populations already partially engaged with health support services rather than those not accessing services.

**Conclusions:** This review of recruitment and retention outcomes with mHealth interventions highlights the need for multimodal recruitment methods. However, significant gaps in effective engagement and retention strategies limit the positive outcomes expected from mHealth interventions.

## BACKGROUND

Addiction disorders and problematic substance use are significant public health problems, requiring a cross-disciplinary and multi-level action approach to effective interventions. The prevalence of addictive disorders and problematic substance use varies by the substance or behaviour of concern and across population groups. For example, international standardized prevalence rates of gambling disorders range from 0.5% to 7.6%, with an average rate across all countries of 2.3%,^1^ while the global prevalence of alcohol use disorders is estimated at 8.6% (95% Confidence Interval (CI) 8.1-9.1) in men and 1.7% (95% CI 1.6-1.9) in women.^2^ Age-standardized prevalence of dependent cannabis use is 3.5%,^3^ and an estimated 22% of the global population smoke tobacco daily and 0.77% use amphetamines daily.^4^ The misuse and abuse of drugs contribute significantly to the global burden of disease. For example, 4.2% (3.7–4.6) of disability-adjusted life years (DALYs) are attributable to alcohol use.

Despite the disease burden and individual harms experienced, and irrespective of the disorder, many people with gambling or substance use disorders are not receiving treatment. Access is a major barrier to care, especially when treatment interventions are primarily delivered face-to-face. ^5, 6^ Financial issues can also be a barrier to treatment; for example, a USA survey of 9000 people with mental health and substance use disorders reported that 15% of respondents did not seek help at all and 17% left treatment early due to financial costs. ^7^ Geographic location has also been found to be a barrier where people living in rural locations have less service provision or fewer choices than their urban counterparts. ^8^ Other barriers to seeking and receiving help include reports of a feeling of shame or stigma as well as a fear of government agencies. ^9^ Cultural appropriateness of services delivered and innate racial bias have also been reported as barriers. ^10^ As a result, significant proportions are without treatment or flexible treatment options. ^11, 12^

mHealth interventions have been suggested as an alternative to overcome many barriers that deter individuals from seeking help. mHealth interventions are typically shorter and found to be more cost-effective, enable immediate treatment access, and have a greater and more diverse reach than analogue interventions. ^13^ Thus, they have the potential to reach a more significant number of those in need of help than traditional intervention models.

### mHealth Tools

mHealth is a catchall term that encompasses and refers to the many different capabilities of mobile phone technology, such as talking, texting, on and offline internet content and sensors within or tethered to mobile phones and applies them to health across the continuum. ^14^

Smartphone applications (apps) are among the more common mHealth tools developed for health interventions. Many have been designed and developed to support self-management and behaviour change for smoking cessation^15^, cardiac rehabilitation, ^16^ healthy lifestyles,^16^ diabetes^17^, Human Immunodeficiency Virus (HIV), ^18^ nutrition, ^19^ mental illness ^20^ and even youth driving. ^21^ mHealth interventions designed and developed using good evidence collectively show promise^22^, and mHealth and app use is growing in other health domains, including other forms of addiction and problematic substance use.^23^ However, relatively few mHealth interventions regarding problem, disordered or harmful gambling have been developed. In contrast, in alcohol and substance misuse, many of the apps that have been developed are not based on empirically supported interventions, have not been empirically tested and are often not readily available.^24^ This lack of evidence can lead to unintended negative consequences, such as delaying help-seeking or promoting information inconsistent with current health advice.^25^ However, apps that have been evaluated, such as those for alcohol - ‘Step Away’, A-CHESS,^26^ Promillekol,^27^ and SMS programmes with and without web-based support or feedback, ^28–30^ have been found effective in reducing substance use long-term. SMS aftercare programmes have also been evaluated in adults discharged from rehabilitation facilities ^31^ and to help adults reduce marijuana use.^32^

### Recruiting hard-to-reach populations

mHealth tools can potentially reach more hard-to-reach populations, such as those with comorbid mental health or substance use disorder and marginalized groups. In smoking cessation trials, retention of participants with mental health or substance use disorders or problematic substance use can be poor compared to other population groups, ^33, 34^ with some trials losing more than two-thirds of their participants at follow-up. In general, people of colour, people from minority groups, and those from lower socioeconomic groups are underrepresented as research participants in clinical trials despite often having an increased disease burden due to socioeconomic determinants of health.^35, 36^

In 2019 it was estimated that 5 billion people worldwide had mobile devices, over half of which were smartphones.^37^ In parallel, mobile technology has been proliferating. New capabilities such as GPS, augmented and virtual reality, wearable and implantable sensors, and biometric authentication are difficult to ignore. These capabilities have highlighted the role mobile devices such as smartphones can play in the addiction intervention space.

### Rationale

This systematic review aimed to identify published peer-reviewed literature on the recruitment and retention of participants in studies of mHealth interventions for people with addiction and/or problematic substance use and to identify successful recruitment strategies. This systematic review focuses on different types of addictive disorders or problematic substance use, such as gambling, tobacco, problematic drinking and the use of these addictive substances.

## METHODS

We conducted a systematic review following the Preferred Reporting Items for Systematic Review and Meta-Analyses (PRISMA) guidelines. ^38^ The review protocol was prospectively registered with PROSPERO (CRD 42021279724).

### Search Strategy

We conducted an electronic search of Medline, Embase, PsychINFO, and Cochrane Central Register of Controlled Trials (CENTRAL). Search terms combined addiction-related terms with mHealth, treatment terms, participant recruitment and retention (see Appendix 1). An electronic search using Google Scholar to check for missed publications (limited to the first 200 results) was also conducted.

We limited the search to peer review literature and English language abstracts or text. Our search was limited to publications after January 1998, when text messaging became mainstream. All searches were conducted up until 02 August 2023.

In addition to our database search, we hand-searched the references of eligible publications for additional references.

### Screening and data extraction

All search results were exported to Rayyan.ai, and duplicates were removed automatically. Titles and abstracts were screened independently by two authors (BK and JCM) against the screening criteria for potential relevance (Table 1). Only results papers were included, although protocol papers, lessons learned, and formative papers could be included as sources of additional information if the results paper was also included. Any disagreements between reviewers were resolved by a third reviewer (GH).

**Table 1.** Study inclusion criteria.

| Criteria | Inclusion | Exclusion |
| --- | --- | --- |
| **Study design** | RCT, cluster-RCT, quasi-RCT, and non-randomized controlled trials | Studies without a control arm<br>Observational studies |
| **Participants** | Adults (18 years or older) with:<br>• Self-reported addiction or problematic substance use / misuse<br>• Diagnosed with addictive disorder (i.e., DSM 304-305, ICD F10-F19)<br>Studies will not be excluded if participants receive concurrent treatment or have comorbid diagnoses. | Mobile phone, social media, and internet addiction |
| **Setting** | No limits are placed on setting |  |
| **Intervention** | Any mHealth intervention to manage addiction or problematic substance use, including: smartphone apps, text or SMS programmes, programmes explicitly using smartphone technology as part of an intervention. | Interventions not primarily delivered via a mobile device |
| **Control** | No limits on the type of control | No control arm |
| **Outcome** | No limits on outcome measured |  |

We retrieved the full-text articles of all relevant articles for further screening by both reviewers. All articles that were not excluded were imported to NVivo for data extraction.

### Study Quality

For all RCTs, the ROB-2 assessment ^39^ was conducted by one reviewer (BK or AO) (See Table 2). For all non-randomized and quasi-randomized studies, ROBINs-I ^40^ was used to evaluate quality (See Table 3). A second reviewer (JM) assessed 20% of included studies. Any disagreements in the scoring between reviewers were resolved by discussion. If disagreements were unresolved by discussion, they were arbitrated (GH).

**Table 2:** Summary of ROB-2 quality assessment for included RCTs.

| Authors, Year | Criteria |  |  |  |  | Overall Bias |
| --- | --- | --- | --- | --- | --- | --- |
|  | Randomization process | Deviations from intended interventions | Missing outcome data | Measurement of the outcome | Selection of the reported result |  |
| Abroms 2014<sup>41</sup> | Low risk | Low risk | Low risk | Low risk | Some concerns | Some concerns |
| Abroms 2017<sup>42</sup> | Low risk | Low risk | Low risk | Low risk | Some concerns | Some concerns |
| Affret 2020<sup>43</sup> | Low risk | High risk | Low risk | Low risk | Low risk | Some concerns |
| Agyapong 2012<sup>28</sup> | Low risk | Low risk | Low risk | Low risk | Some concerns | Some concerns |
| Agyapong 2018<sup>44</sup> | Low risk | Low risk | Low risk | Low risk | Some concerns | Some concerns |
| Aigner 2017<sup>45</sup> | Some concerns | Low risk | Low risk | Low risk | Some concerns | Some concerns |
| Asayut 2022 | Low risk | Low risk | Some concerns | Low risk | Some concerns | Some concerns |
| Bindhim 2018<sup>15</sup> | Low risk | Low risk | Low risk | Low risk | Some concerns | Some concerns |
| Bricker 2014<sup>46</sup> | Low risk | Low risk | Low risk | Low risk | Low risk | Low risk |
| Bricker 2020<sup>47</sup> | Low risk | Low risk | Low risk | Low risk | Some concerns | Some concerns |
| Chen 2020<sup>48</sup> | Low risk | Low risk | Low risk | Low risk | Low risk | Low risk |
| Cheung 2015<sup>49</sup> | High risk | Some concerns | Some concerns | Low risk | Some concerns | High risk |
| Demartini 2018<sup>50</sup> | Low risk | Some concerns | Low risk | Low risk | Low risk | Some concerns |
| Destasio 2018<sup>51</sup> | Some concerns | Some concerns | Low risk | Low risk | Some concerns | Some concerns |
| Farren 2022 | Some concerns | Some concerns | Some concerns | Some concerns | Some concerns | Some concerns |
| Forinash 2018<sup>52</sup> | Low risk | Some concerns | Low risk | Low risk | Some concerns | Some concerns |
| García Pazo 2021 | Some concerns | Some concerns | Some concerns | Low risk | Some concerns | Some concerns |
| Goldenhersch 2020<sup>53</sup> | Low risk | Some concerns | Low risk | Low risk | High risk | High risk |
| Graham 2020<sup>54</sup> | Low risk | Low risk | Low risk | Low risk | Low risk | Low risk |
| Graham 2021<sup>55</sup> | Low risk | Low risk | Low risk | Low risk | Low risk | Low risk |
| Gustafson 2014<sup>56</sup> | Low risk | Some concerns | Low risk | Low risk | Low risk | Low risk |
| Haug 2017<sup>57</sup> | Low risk | Low risk | Some concerns | Low risk | Low risk | Low risk |
| Hébert 2020<sup>58</sup> | Low risk | Some concerns | Low risk | Low risk | Some concerns | Some concerns |
|  | Randomization process | Deviations from intended interventions | Missing outcome data | Measurement of the outcome | Selection of the reported result |  |
| Hicks 2017<sup>59</sup> | Some concerns | High risk | Low risk | Low risk | High risk | High risk |
| Hides 2018<sup>60</sup> | Some concerns | Some concerns | Low risk | Low risk | Low risk | Some concerns |
| Hoepfner 2017<sup>61</sup> | High risk | Some concerns | Low risk | Low risk | Some concerns | High risk |
| Keoleian 2013<sup>62</sup> | High risk | Some concerns | Low risk | Low risk | Some concerns | High risk |
| Liang 2018<sup>63</sup> | Some concerns | Some concerns | Low risk | Low risk | Some concerns | Some concerns |
| Lucht 2021<sup>64</sup> | Low risk | Some concerns | Low risk | Low risk | Some concerns | Some concerns |
| Masaki 2020<sup>65</sup> | Low risk | Low risk | Low risk | Low risk | Low risk | Low risk |
| Mason 2014<sup>66</sup> | Some concerns | Some concerns | Low risk | Low risk | High risk | Some concerns |
| Mason 2018b<sup>67</sup> | Low risk | Some concerns | Low risk | Low risk | Some concerns | Some concerns |
| McTavish 2012<sup>68</sup> | High risk | High risk | High risk | Some concerns | High risk | High risk |
| Muench 2017<sup>69</sup> | Low risk | Some concerns | Some concerns | Low risk | Low risk | Some concerns |
| Mussener 2016<sup>70</sup> | Low risk | Some concerns | Low risk | Low risk | Low risk | Low risk |
| Pechmann 2017<sup>71</sup> | Some concerns | Some concerns | Low risk | Low risk | Low risk | Some concerns |
| Reback 2019<sup>72</sup> | Some concerns | Some concerns | Some concerns | Some concerns | Low risk | Some concerns |
| Rodda 2018<sup>73</sup> | Low risk | Low risk | Low risk | Low risk | Some concerns | Low risk |
| Schlam 2020<sup>74</sup> | Low risk | Low risk | Low risk | Low risk | Some concerns | Low risk |
| Scott 2020<sup>75</sup> | Some concerns | Some concerns | Low risk | Some concerns | Low risk | Some concerns |
| So 2020<sup>76</sup> | Low risk | Low risk | Low risk | Low risk | Some concerns | Low risk |
| Sridharan 2019<sup>77</sup> | Low risk | Low risk | Low risk | Some concerns | Low risk | Low risk |
| Villardaga 2020<sup>78</sup> | Some concerns | Some concerns | Low risk | Low risk | Low risk | Some concerns |
| Webb 2020<sup>79</sup> | Low risk | Low risk | Low risk | Low risk | Low risk | Low risk |
| Whittaker 2011<sup>80</sup> | Low risk | Low risk | Low risk | Low risk | Low risk | Low risk |
| Witkiewitz 2014<sup>81</sup> | Some concerns | Some concerns | Low risk | Low risk | Low risk | Some concerns |
| Xu 2021<sup>82</sup> | Low risk | Low risk | Low risk | Low risk | Low risk | Low risk |
| Ybarra 2013<sup>83</sup> | Low risk | Low risk | Some concerns | Low risk | Low risk | Low risk |

**Table 3:** Summary of ROBIN-S quality assessment for included non-RCTs.

| Authors, Year | Criteria |  |  |  |  |  |  | Overall bias |
| --- | --- | --- | --- | --- | --- | --- | --- | --- |
|  | Confounding | Selection of participants into the study | Classification of interventions | Deviations from intended interventions | Missing data | Measurement of outcomes | Selection of the reported result |  |
| Chen 2019<sup>84</sup> | Moderate | Low | Low | Low | Low | Low | Low | Low |
| Eiler 2020<sup>85</sup> | Low | Low | Low | Moderate | Low | Low | Low | Low |
| Rajani 2021<sup>86</sup> | Low | Low | Low | Low | Low | Low | Low | Low |
| Vilaplana 2014<sup>87</sup> | Low | Low | Low | Low | Moderate | Moderate | Serious | Moderate |
| Gonzalez and Dulin 2015<sup>88</sup> | Low | Low | Low | Low | Moderate | Low | Low | Low |

### Data synthesis

We summarised study information and conducted a narrative review and qualitative literature synthesis to summarise the findings across studies. We used descriptive statistics to summarise the study and participant characteristics where appropriate.

The following outcomes were evaluated where possible:

- Types of recruitment strategies used
- Effectiveness of different recruitment strategies
- Population groups targeted
- Effectiveness of recruitment strategies for different population groups and different disorders
- Explanation for differential recruitment

We also compared different recruitment methods on recruitment and retention numbers and how effective strategies were with different population groups.

## RESULTS

### Study selection

The electronic search results in 2135 papers. After duplicates were removed, 1654 papers were screened for eligibility. After initial screening by title and abstract, 1550 papers were excluded, and 104 were included, including eight additional results papers identified from protocol papers meeting criteria. After full-text screening, there were 60 relevant papers (Figure 1). Of these papers, 53 were primary analyses, five secondary analyses and two protocols (Table 4). Study designs for the primary analyses included 45 randomized controlled trials, one pseudo-randomized trial, five non-randomized trials, and one cluster randomized controlled trial.

**Figure 1:**
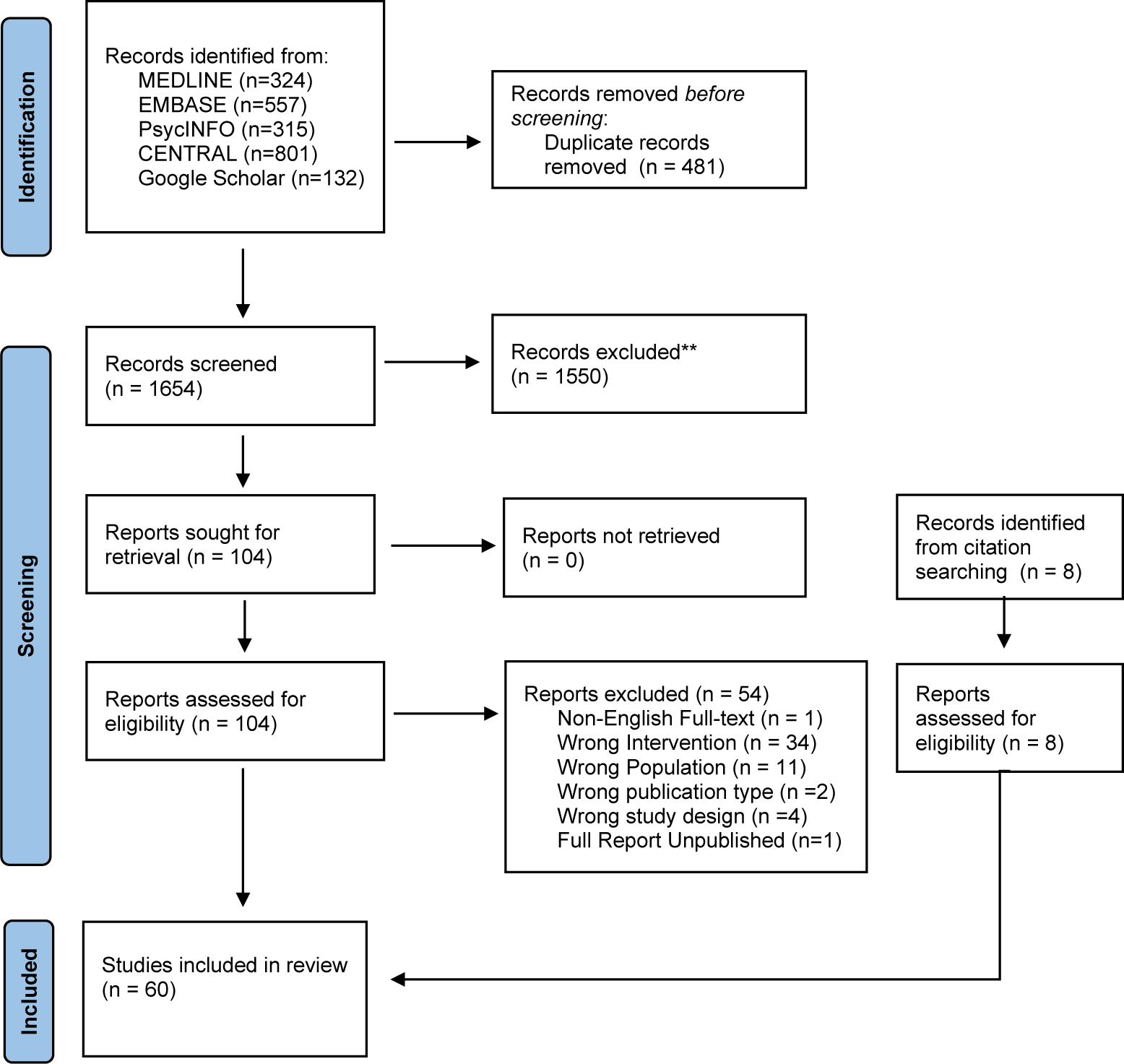
PRISMA Flow diagram.

**Table 4:** Overview of included studies, characteristics and recruitment.

| Authors, Year | Study Design | Country | Topic | Population | Intervention | Recruitment |  |  |
| --- | --- | --- | --- | --- | --- | --- | --- | --- |
|  |  |  |  |  |  | Sample Size^ |  | Methods^ |
|  |  |  |  |  |  | Target | Actual |  |
| Abroms et al., 2014<sup>38</sup> | RCT | US | Smoking | Adult Smokers (18+ y/o) | Text messaging program (Text2Quit) | -- | -- | No Information (--) |
| Abroms et al., 2017<sup>39</sup> | RCT | US | Smoking | Pregnant women | Text messaging program (Text2Quit) | -- | -- | -- |
| Affret et al., 2020<sup>40</sup> | RCT | France | Smoking | Adult Smokers | Web & Mobile application (e-Tabac Info Service (e-TIS)) | -- | -- | -- |
| Agyapong et al., 2012<sup>25</sup> | RCT | Ireland | Alcohol | Adults with Major Depressive Disorder and Alcohol Dependency | Text message program | -- | -- | -- |
| Agyapong et al., 2018<sup>41</sup> | RCT | Canada | Alcohol | Adults with Alcohol Use Disorder, final week of addiction treatment program | Text message program | 60 | 59 | In-person (Addiction treatment program at Rehabilitation Centre) |
| Aigner et al., 2017<sup>42</sup> | RCT | US | Smoking | Smokers with HIV+ status | Cell-phone-based counselling sessions | -- | -- | -- |
| Asayut et al., 2022<sup>89</sup> | RCT | Thailand | Smoking | Thai Smokers | The “PharmQuit” app | -- | -- | In-person (invitation by pharmacy students, community pharmacists, health care providers, and health care volunteers) |
| Bindhim et al., 2018<sup>12</sup> | RCT | US Australia SingaporeUK | Smoking | Adult Smokers (18+ y/o) | Mobile application (Smartphone Smoking Cessation App (SSC APP)) | -- | -- | -- |
|  |  |  |  |  |  | Sample Size^ |  | Methods^ |
|  |  |  |  |  |  | Target | Actual |  |
| Bricker et al., 2014 <sup>43</sup> | RCT | US | Smoking | Adult Smokers (18+ y/o) | Mobile application (SmartQuit) | 196 | 196 | Online (Facebook advertisements, website advertisements, and search engine ads)<br>Traditional (television, newspaper and radio advertisements) |
| Bricker et al., 2020 <sup>44</sup> | RCT | US | Smoking | Adult Smokers (18+ y/o) | Mobile application (iCanQuit) | 2500 | 2503 | Online (Facebook advertisements, survey sampling company, search engine advertisements) |
| Cambon et al., 2017 <sup>85*</sup> | RCT - Protocol | France | Smoking | Adult Smokers (18+ y/o) | Web & Mobile application (e-Tabac Info Service (e-TIS)) | 3000 | NA | Online (advertisement on French Mandatory National Health Insurance website) |
| Chen et al., 2019 <sup>81</sup> | RCT | China | Alcohol | Adults 20-50 y/o, diagnosed with Alcohol Dependence | CBT on the WeChat platform | -- | -- | -- |
| Chen et al., 2020 <sup>45</sup> | Non-RCT | China | Smoking | Adult Smokers (25-44 y/o) | CBT on the WeChat platform (Smoking Cessation Intervention (SCAMPI)) | -- | -- | -- |
| Cheung et al., 2015 <sup>46</sup> | Cluster-RCT | China | Smoking | Adult Smokers (18+ y/o) | WhatsApp or Facebook online social group | -- | -- | -- |
| Demartini et al., 2018 <sup>47</sup> | RCT | US | Alcohol | Recent drinking episode in past year (1+) | Text messaging program | -- | -- | -- |
| Destasio et al., 2018 <sup>48</sup> | RCT | US | Smoking | General Public not diagnosed with substance use, psychiatric or neurological disorder | Text messaging program | -- | -- | -- |
|  |  |  |  |  |  | Sample Size^ |  | Methods^ |
|  |  |  |  |  |  | Target | Actual |  |
| Eiler et al., 2020<sup>82</sup> | Non-RCT | Germany | Smoking | Smokers | Mobile application (Approach-Avoidance Task (app-AAT)) | -- | -- | -- |
| Farren et al., 2022<sup>90</sup> | RCT | Ireland | Alcohol | Adults 18-70 with Alcohol Use Disorder as the primary disorder, who were inpatients completing therapeutic programmes. | Mobile application "UControlDrink". The app incorporates daily supportive text messaging and C-CBT | -- | -- | Participants were recruited in-person from St. Patrick's University Hospital, Dublin. |
| Forinash et al., 2018<sup>49</sup> | RCT | US | Smoking | Pregnant women | Text messaging program | 60 | 49 | In-person (Maternal Fetal Care Center) |
| Garcia-Pazo et al., 2021<sup>91</sup> | Pseudo-Randomized Clinical Trial | Spain | Smoking | Smokers admitted to a public hospital in the Migjorn health sector in the Balearic Islands. | Mobile application "NoFumo+". | -- | -- | In-person. Interest was gauged at admission to hospital and recruitment was completed within 48 hours of first contact. |
| Glass et al., 2017<sup>86</sup> | RCT - Secondary Analysis | US | Alcohol | Adults diagnosed with Alcohol Dependency (18+) | Mobile application (Addiction-Comprehensive Health Enhance Support System (A-CHES)) | -- | -- | -- |
| Goldenhersch et al., 2020<sup>50</sup> | RCT | Argentina | Smoking | Adult Smokers (24-65 y/o) | Mobile application | -- | -- | -- |
| Gonzalez & Dulin, 2015<sup>84</sup> | Non-randomized controlled trial | US | Alcohol | Adults diagnosed with Alcohol Use Disorder (18-45 y/o) | Mobile application (Location-Based Monitoring and Intervention for Alcohol Use Disorders (LBMI-A)) | -- | -- | -- |
| Graham et al., 2020<sup>51</sup> | RCT | US | Smoking | Adult Smokers (18+) | Web program (EX) and Text message program | -- | -- | -- |
| Graham 2021<sup>52</sup> | RCT | US | Vaping | Adolescents e-cigarette users (18-24 y/o) | Text messaging program (This is Quitting (TIQ)) | 2600 | 2588 | Online (advertisements on Facebook and Twitter) |
|  |  |  |  |  |  | Sample Size^ |  | Methods^ |
|  |  |  |  |  |  | Target | Actual |  |
| Gustafson et al., 2014 <sup>53</sup> | RCT | US | Alcohol | Adults (18+ y/o) | Mobile application (Addiction-Comprehensive Health Enhance Support System (A-CHES)) | 350 | 349 | In-person (Residential treatment programs) |
| Han et al., 2018 <sup>87</sup> | RCT - Secondary analysis | China | Substance Use | Adults diagnosed with heroin dependence or amphetamine-type stimulants (ATS) dependence | Mobile application (S-Health) | -- | -- | -- |
| Haug et al., 2017 <sup>54</sup> | Cluster-RCT | Switzerland | Smoking & Alcohol | Smokers | Web and text messaging program | -- | -- | -- |
| Hébert et al., 2020 <sup>55</sup> | RCT | US | Smoking | Adult Smokers (18+ y/o) | Mobile application (Smart-T2) | -- | -- | -- |
| Hicks et al., 2017 <sup>56</sup> | RCT | US | Smoking | Adult Smokers with diagnosed PTSD (18-70 y/o) | Mobile contingency management smoking cessation counselling and medication as and Mobile application (Stay Quit Coach) | -- | -- | -- |
| Hides et al., 2018 <sup>57</sup> | RCT | Australia | Alcohol | Adolescents with monthly alcohol use (16-25 y/o) | Mobile application (Ray's Night Out) | -- | -- | -- |
| Hoepfner et al., 2017 <sup>58</sup> | RCT | US | Smoking | Adult Smokers (18+ y/o) | Text messaging program (Text2Quit) | -- | -- | -- |
| Keoleian et al., 2013 <sup>88</sup> | RCT | US | Substance Use | Methamphetamine users seeking treatment | Text messaging program | -- | -- | -- |
| Liang et al., 2018 <sup>60</sup> | RCT | China | Substance Use | Adults, used heroin or other substance use in past 30 days | Mobile application (S-Health) | -- | -- | -- |
| Lucht et al., 2021 <sup>61</sup> | RCT | Germany | Alcohol | Adults diagnosed with alcohol dependence, ongoing inpatient alcohol detoxification (18+ y/o) | Text messaging program (Continuity of care among alcohol-dependent patients (CAPS)) | 462 | 463 | In-person (Psychiatric hospitals) |
|  |  |  |  |  |  | Sample Size^ |  | Methods^ |
|  |  |  |  |  |  | Target | Actual |  |
| Luk et al., 2019* <sup>89</sup> | Cluster-RCT - Protocol | China | Smoking | Adults Smokers | WhatsApp chat-based support | 1172 | NA | NA |
| Masaki et al., 2020 <sup>62</sup> | RCT | Japan | Smoking | Adults diagnosed with nicotine dependence | Mobile application (Cure App Smoking Cessation (CASC)) | 580 | 584 | In-person (Smoking cessation clinics) |
| Mason et al., 2014 <sup>63</sup> | RCT | US | Alcohol | Individuals diagnosed with problem drinking | Text messaging program (TROPO) | -- | -- | -- |
| Mason et al., 2018a <sup>90</sup> | RCT - Secondary analysis | US | Substance Use | Adolescents diagnosed with cannabis use disorder (18-25 y/o) | Text messaging program (Peer Network Counseling (PNC-txt)) | -- | -- | -- |
| Mason et al., 2018b <sup>64</sup> | RCT | US | Substance Use | Adolescents diagnosed with cannabis use disorder (18-25 y/o) | Text messaging program (Peer Network Counseling (PNC-txt)) | -- | -- | -- |
| Mason 2020<sup>91</sup> | RCT - Secondary analysis | US | Substance Use | Adolescents diagnosed with cannabis use disorder (18-25 y/o) | Text messaging program (Peer Network Counseling (PNC-txt)) | -- | -- | -- |
| Mctavish et al., 2012 <sup>65</sup> | RCT | US | Alcohol | Adults (18+ y/o) | Mobile application (Addiction-Comprehensive Health Enhance Support System (A-CHES)) | -- | -- | -- |
| Muench et al., 2017 <sup>66</sup> | RCT | US | Alcohol | Adults consuming 13-15 standard drinks/week (21-65 y/o) | Text messaging program | -- | -- | -- |
| Mussener et al., 2016 <sup>67</sup> | RCT | Sweden | Smoking | Smokers | Text messaging program (Nicotine Exit (NEXit)) | 1354 | 1590 | In-person (Student health care centres) |
| Pechmann et al., 2017<sup>68</sup> | RCT | US | Smoking | Adult Smokers (15-59 y/o) | Twitter peer support group | 160 | 160 | Online (Google advertisements using keywords fo quitting and support) |
|  |  |  |  |  |  | Sample Size^ |  | Methods^ |
|  |  |  |  |  |  | Target | Actual |  |
| Rajani et al., 2021<sup>83</sup> | Non-RCT | UK | Smoking | Adult Smokers (18+ y/o) | Mobile application (Quit Genius) | 140 | 116 | Online (social media)<br>Traditional (posters displayed across public places in London) |
| Reback et al., 2019<sup>69</sup> | RCT | US | Substance Use | Adults with methamphetamine use and reported condomless anal intercourse | Text messaging program | -- | -- | -- |
| Rodda et al., 2018<sup>73</sup> | RCT | Australia | Gambling | Individuals engaged with 1+ service by Gambling help online | Text messaging program | -- | -- | -- |
| Schlam & Baker, 2020<sup>71</sup> | RCT | US | Smoking | Individuals with e-cigarette use | Mobile game applications (arcade, puzzle, word, board, card, tower defense and running games) | -- | -- | -- |
| Scott et al., 2020<sup>72</sup> | RCT | US | Substance Use | Individuals diagnosed with substance use disorder) | Mobile application (Addiction-Comprehensive Health Enhance Support System (A-CHES)) | -- | -- | -- |
| So et al., 2020<sup>73</sup> | RCT | Japan | Gambling | Adults with Problem Gambling | Text messaging program (GAMBOT) | 198 | 197 | Online (Search engine advertisements for users searching for helpful information to stop gambling) |
| Sridharan et al., 2019<sup>74</sup> | RCT | US | Smoking | Adults Smokers (18+) | Mobile application (SmartQuit) and web-de;overed growth mindset intervention | 300 | 398 | Online (Facebook advertisements and internet survey panel) |
|  |  |  |  |  |  | Sample Size^ |  | Methods^ |
|  |  |  |  |  |  | Target | Actual |  |
| Tamí-maury et al. 2013 <sup>92</sup> | RCT - Secondary Analyses | US | Smoking | Adult Smokers with HIV+ status (18+ y/o) | Cell-phone-based counselling sessions | -- | -- | -- |
| Vilaplana et al., 2014 <sup>37</sup> | Non-randomized controlled trial | Spain | Smoking | Patients at Smoking Cessation Program of Santa Maria Hospital | Web-based application (Smoker Patient Control (S-PC)) | -- | -- | -- |
| Vilardaga 2020<sup>75</sup> | RCT | US | Smoking | Adult Smokers diagnosed with either Schizophrenia, Schizoaffective, Bipolar, or recurrent major depressive disorder (18+ y/o) | Mobile applications (Learn to Quit & QuitGuide) | 90 | 92 | **In-person** (coordinating with primary care clinics, collaboration with smoking cessation programs and mental health clinics)<br>**Online** (electronic health records, patient health portal invitations) |
| Webb et al., 2020 <sup>76</sup> | RCT | UK | Smoking | Adult Smokers (18+ y/o) | Mobile application (Quit Genius) | 388 | 556 | In-person (Primary care practices) |
| Whittaker et al., 2011 <sup>(77)</sup> | RCT | NZ | Smoking | Young Māori Smokers (16+ y/o) | Video-based text messaging program | 1300 | 226 | **Online** (Online magazine, Internet and mobile phone advertisements)<br>**Traditional** (Advertisements to radio, paper-based magazines, Māori-specific media, local and national newspapers, and media releases) |
|  |  |  |  |  |  | Sample Size^ |  | Methods^ |
|  |  |  |  |  |  | Target | Actual |  |
| Witkiewitz et al., 2014<sup>78</sup> | RCT | US | Smoking & Alcohol | College students with concurrent smoking and drinking and/or recent heavy drinking episode(s) | Mobile application (Brief Alcohol Screening and Intervention for College Students (BASICS)) | -- | -- | -- |
| Xu et al., 2021<sup>79</sup> | RCT | China | Substance Use | Adults diagnosed with substance use disorder (20-50 y/o) | Web and Mobile application (Community-based addiction rehabilitation electronic system (CAREs)) and community-based rehabilitation | -- | -- | -- |
| Ybarra et al., 2013<sup>80</sup> | RCT | US | Smoking | Adolescent Smokers (18-25 y/o) | Text messaging program (Text Care) and Text peer support program (Text Buddy) | -- | -- | -- |

\*Study Protocol
RCT = Randomised Control Trial
^Blanks are for papers that did not report their sample size numbers and/or method. **Target** number of randomized participants according to either the power calculations or stated intentions of the study authors. **Actual** number of randomized participants included in the study.
NA=Not Available

### Study characteristics

The included studies primarily comprised primary analyses (N=53) ^15, 28, 41–88 89 90 91^, secondary analyses (N=5),^92–96^ and study protocols (N=2).^97, 98^ The primary analyses were mostly RTCs (N=45),^15, 28, 41–47, 50–56, 58–84 89, 90^ pseudo-RCTs (N=1),^91^ cluster RCTs (N=2),^49, 97^ and non-RCTs (N=5).^48, 85–88^ Secondary analyses were all based on RCTs, and the study protocols were based on an RCT and cluster RCT design each.

Most of the included studies were concerned with smoking cessation (N=33), ^15, 41–43, 45–49, 51-54, 58, 59, 61, 65, 70, 71, 74, 77–80, 83, 85–87, 89, 91, 92, 97, 98^ of which 30 were primary analyses, one secondary analysis, and two study protocols. Nine studies were concerned with problematic substance use (i.e., heroin, cannabis), ^62, 63, 67, 72, 75, 82, 93–95^ of which six were primary analyses and three secondary analyses. Twelve studies were concerned with alcohol use, ^28, 44, 50, 56, 60, 64, 66, 68, 69, 84, 88, 96^ of which 11 were primary analyses and one a secondary analysis. Two studies were concerned with co-occurring alcohol use and smoking, ^57, 81^ all of which were primary analyses. Two primary analyses were found of interventions for problem gambling.^73 75^ One study was concerned with vaping cessation ^55^ as the primary analysis.

More than half of the included studies were from the United States (US) (n=32).^41, 42, 45–47, 50-52, 54–56, 58, 59, 61, 62, 66–69, 71, 72, 74, 75, 77, 78, 81, 83, 88, 92–94, 96^ Twelve studies were from Europe (including two from the United Kingdom (UK)),^28, 43, 57, 64, 70, 79, 85–87, 98^ ten from Asia,^48, 49, 63, 65, 76, 82, 84, 95, 97 89^ and one from South America.^53^ Two studies were from Australia,^60, 73^ one from New Zealand (NZ)^80^ and one from Canada.^44^ The remaining study was a multi-site study that included participants from the USA, Australia, Singapore, and the UK.^15^

The most common mHealth intervention was text message programs (n=20)^28, 41, 42, 44, 50–52, 55, 61, 62, 64, 66, 67, 69, 70, 72, 73, 76, 93, 94^ and mobile applications (n=25)^15, 46, 47, 53, 56, 58–60, 63, 65, 68, 74, 75, 77–79, 81, 85, 86, 88, 95, 96 89–91^ followed by social media groups (Facebook, Twitter, WhatsApp, and WeChat) (n=5),^48, 49, 71, 84, 97^ web and mobile applications (n=3),^43, 82, 98^ web and text messaging program (n=2),^54, 57^ video-based text message program (n=1)^80^, text message and text peer support program (n=-1),^83^ web application (n=1)^87^ and cell-phone based counselling sessions (n=2).^45, 92^ The number of participants ranged from 5 to 2806. Twenty-one studies had less than 100 participants, twelve were 100-199 participants, and fourteen had 200-499 participants. Eleven studies had more than 500 participants, of which six had more than 1000 participants (see Table 4).

### Risk of Bias

The quality and risk of bias assessment for the RCTs showed that the majority of papers were deemed as ‘low risk’ or ‘some concerns’ for overall bias. The main areas of concern were limited information on the selection of the reported result and potential deviations for intended interventions. The few RCTs deemed ‘high risk’ were mainly for providing little information on the selection of the reported result, randomization process, or concealment process.

For the non-RCTs, the quality and risk of bias assessment revealed the papers as ‘low’ for overall bias except for Vilaplana,^42^ deemed as ‘moderate’. Within each criteria domain, each criterion was mainly deemed as ‘low’ for the papers except for the missing data, measurement of outcomes, confounding, and deviations from intended intervention domains. The moderate rating for Vilaplana was due to little information on missing data, concealment among outcome assessors, selection of the reported result, and the administration of the study instruments. ^42^

### Study Findings

Only 17 studies reported on their recruitment targets, of which 2 were protocols (see Table 4). Eleven studies met their recruitment target ^47, 64, 65, 70, 71, 76–79, 86, 97^ and six did not meet their target.^44, 52, 55, 56, 76, 86^

The overall retention rates as follow-up according to the timeline reported from the included studies (23.1%-100%) (n=32) are summarised in Table 5. According to Dettori (2011),^100^ retention rates of 95% or greater represent little bias (in green), 80-95% show some bias (in yellow), and less than 80% represent serious bias (in red). Only three studies retained recruitment above 95% at their longest follow-up period.^67, 75, 78^ Two studies retained 95% at the first follow-up but fell below 95% at subsequent follow-up.^60, 94^ Thirteen studies ^41, 42, 49, 53, 55, 56, 58, 60, 80, 94^ retained less than 80% of participants at their final follow-up, including four studies that retained less than half of their participants at follow-up (see Table 6).

**Table 5:**
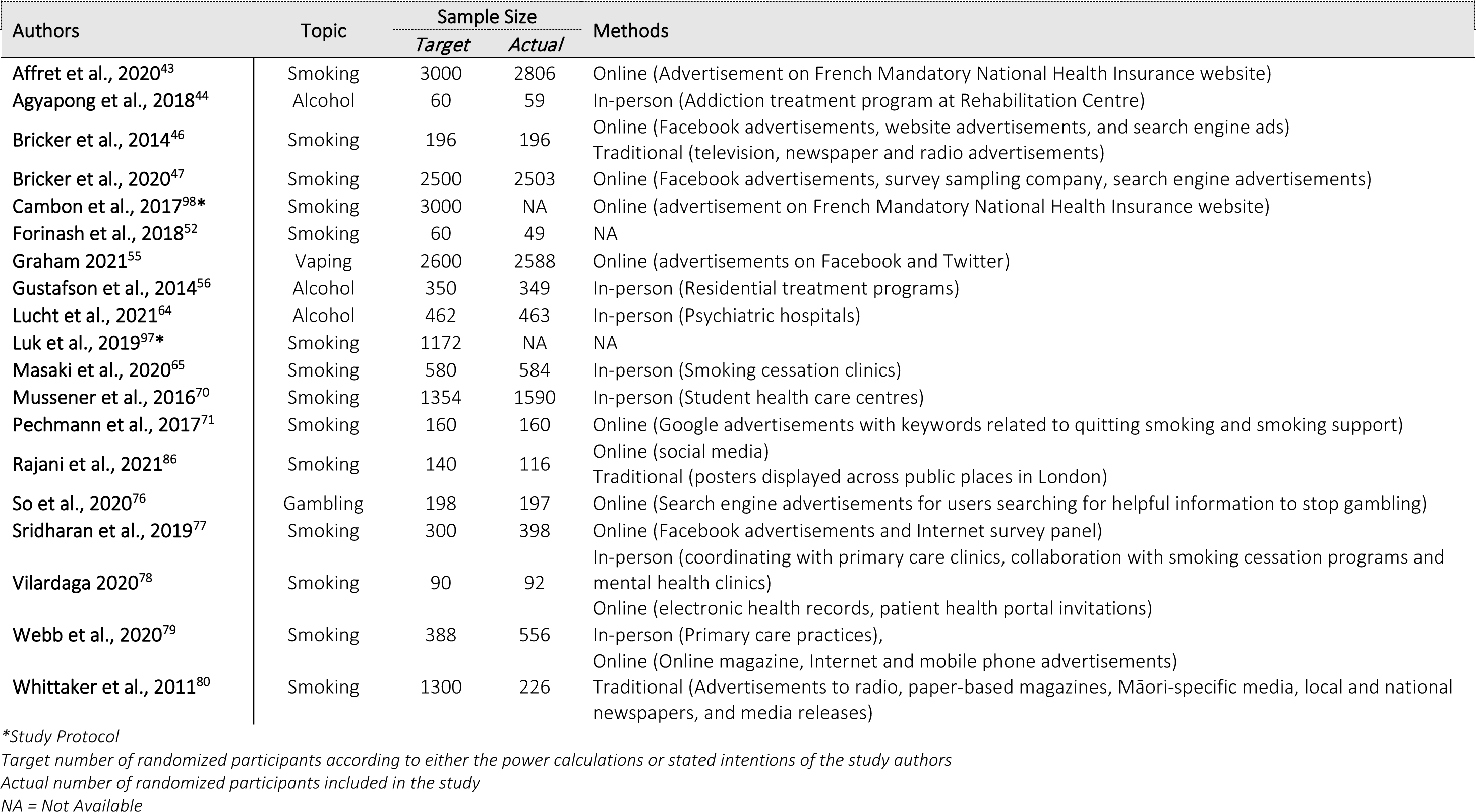
Overview of type of problem, sample size target and actual numbers and recruitment method.

**Table 6:**
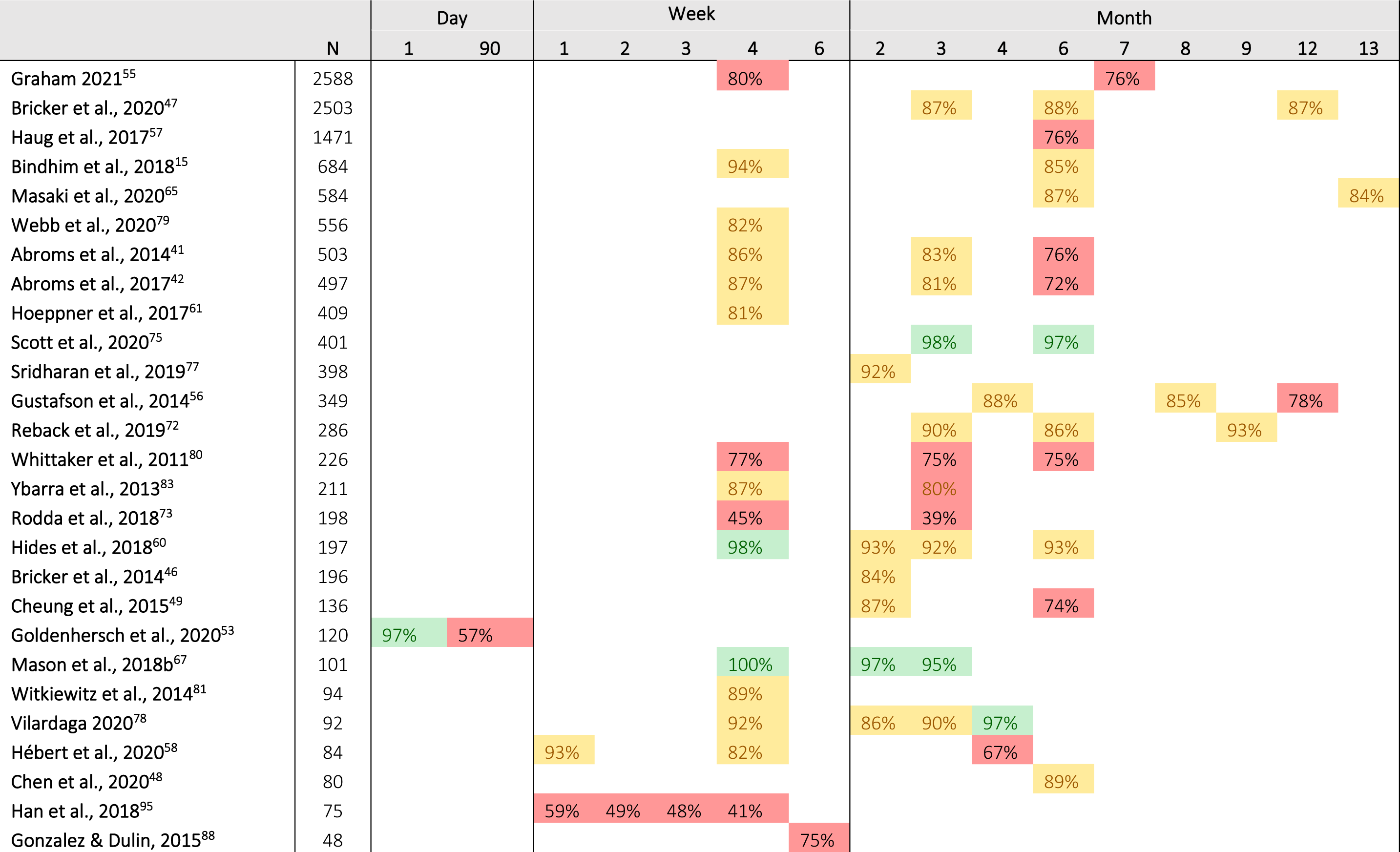
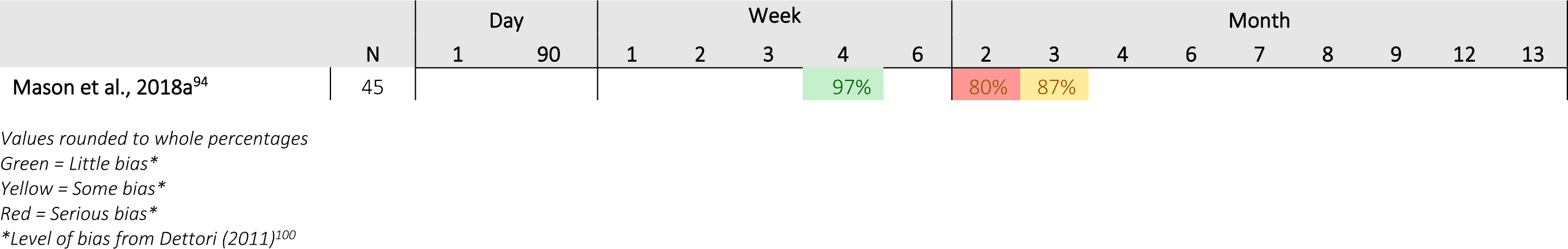
Follow-up rates from the largest to smallest final sample size.

In studies with high retention of participants (at least one follow-up rate ≥95% after Day 1), researchers appeared to use a combination of in-person (e.g., coordinating with healthcare clinics), traditional (e.g. print flyers and radio advertising) and online recruitment (e.g., digital signs, electronic health records) methods, rather than relying solely on one type of recruitment method.^60, 67, 75, 78^ These studies also predominately involved populations already actively engaged in an existing service (e.g., treatment, health services) or easily accessible to the researchers (e.g., university students). As universities commonly conduct this research, these populations are usually easier to recruit and engage.

Follow-up methods used by studies with high retention included screening assessments conducted in person rather than online screening methods. ^60, 67, 75, 78^ Using in-person screening methods could result in the screening assessment being conducted more rigorously and capturing participants more likely to adhere to a mHealth intervention.

In studies with the lowest retention (at least one follow-up rate ≥70% after Day 1), researchers appeared to predominately rely on one primary recruitment method, explicitly using in-person recruitment through healthcare services or treatment programs.^53, 58, 73, 95^ The mHealth interventions for these studies commonly include applications with mobile phone-based ecological momentary assessment (EMA) features. Authors of one of the studies have suggested that combining pre-intervention education with the EMA technology could increase adherence to the intervention among participants.^95^ Screening among the studies with low retention reported that they conducted the screening online, which may have resulted in participants who were less likely to adhere to the study being included compared to in-person screening.

### Problematic Substance Use (excluding alcohol)

Nine studies evaluated digital interventions for problematic substance use, including cannabis, opioids, and stimulants. ^62, 63, 67, 72, 75, 82, 93–95^ Recruitment methods included traditional media, such as flyers, radio, and print media, as well as online media, such as Craigslist, dating apps, and social media advertising. Five studies also conducted recruitment through treatment providers. The studies generally relied on multiple recruitment methods, primarily when recruiting hard-to-reach populations (i.e., Men Who Have Sex with Men (MSM) using methamphetamine vs. college students).^72, 94^ These studies included participants recruited from college student participant pools, clinical settings, and public settings. None of these studies reported a recruitment target. The actual recruitment numbers ranged from 5 participants to 401 participants. Four of the studies only included participants who met the Diagnostic Statistical Manual of Mental Disorders (DSM) criteria for substance use disorder,^67, 72, 75, 82, 94, 95^ two studies included participants who had used substances in the previous 24 hours or 30 days, and one used the addiction severity index.^63^. Two studies described the relationship of the recruiter to participants; one described it as generally by research staff attending treatment agencies with patients^75^, the other hired social workers to conduct trainings.^82^

Participant retention was relatively high across all seven studies, with more than 80% of participants retained in all seven studies. Reasons for drop-out included arrests and withdrawal from the study. In three studies, participants were compensated for completing follow-up measures .^67, 72, 94^ Compensation ranged from USD 25 to USD 50 per follow-up and up to USD 150 over the course of a study. Retention was also maintained through community outreach^72,77^ and treatment providers. ^63^ Follow-up ranged from 4 weeks (i.e., following intervention) to 9 months.

### Alcohol

Thirteen studies evaluated digital interventions for alcohol use, including two interventions that combined alcohol and tobacco interventions.^28, 44, 50, 56, 57, 60, 64, 66, 68, 69, 81, 84, 88, 96^ Seven interventions targeted participants meeting formal diagnostic criteria (i.e., Diagnostic and Statistical Manual of Mental Disorders), including three for alcohol use disorder. Ten studies recruited participants through treatment providers, including hospitals, outpatient clinics, and rehabilitation facilities,^28, 44, 50, 56, 64, 68, 84, 96 90, 91^ and one study recruited participants from schools.^57^ The remaining studies recruited through traditional and online media,^60, 69, 81^ including one study that recruited through online help-seeking resources.^69^ Three studies reported meeting their recruitment targets,^44, 56, 64^ one study ^64^ met their target while two did not, although they missed achieving their target by one participant.^44, 56^ Recruitment numbers ranged from 15 to 1471 participants. Participants were recruited over five months to 33 months. Of the included papers discussing the recruiter role, recruiters were typically study therapists, research assistants or project coordinators attending the primary setting (health clinic or school, for example) of their potential participants to discuss and enable them to take part.^50, 56, 57, 64^

Most of the studies from clinical settings reported high retention of participants (i.e., follow-up for more than 90% of participants, ^28, 50, 60, 66, 84^ four studies reported retention under 90% at 89%,^81^ 75%,^88^ 73% ^44^ and 78%.^56^ Retention of 76% was reported for the school setting.^57^ The remaining studies reported retention equal to or above 90%.^28, 50, 60, 66, 84^. Follow-up ranged from 4 weeks to 12 months. In four studies, participants were compensated for completing surveys, ^57, 69, 81, 88^ which ranged from 10 Swiss Francs (USD$ 10.23) to up to USD 168 for completing the study.

### Gambling

Two of the included studies were digital interventions for problem gambling.^73, 76^ Both were text-based interventions using SMS or messenger apps as the mode of delivery. The two studies used different recruitment methods: one recruited through traditional methods via a gambling helpline, while the other used online advertisement. The recruitment duration was 5 and 12 months. Only one of the studies indicated a recruitment target; the target of 198 participants was exceeded, although only 197 participants were analyzed.^76^ Both studies recruited more than 200 participants. The two studies differed in the stringency of the eligibility criteria. In Scott,^76^ participants were eligible if their problem gambling severity index (PGSI) score of three or greater. They were excluded if they were receiving face-to-face support for their gambling problems. In contrast, participants were included in Rodda^73^ if they were engaged with at least one service offered by the Gambling Helpline. Retention rates also varied between the two studies: Rodda^73^ reported 50% drop-out while Scott ^76^ reported post-intervention assessment for 91-97% of participants analyzed; however, the study follow-up period was only four weeks compared to 12 weeks.

### Smoking Cessation: Hard-to-Reach Populations

Only six studies recruited specific population groups or were targeted for a specific population group, including smokers with HIV-positive status,^45^ pregnant women,^42, 52^ smokers with a diagnosis of PTSD, schizophrenia, schizoaffective disorder, or mood disorders,^59, 78^ and young Māori smokers.^80^ Eligibility criteria included a minimum number of cigarettes per day for most studies; however, in the trials targeting pregnant women, any cigarette consumption was sufficient to meet eligibility. Three of the studies recruited participants through clinical settings,^45, 52, 78^ with two of those using participants already receiving care at the associating clinic. ^45, 52^ One study recruited participants solely through online media^41^, and the last study used traditional and online media.^80^ One study did not provide recruitment information.^59^ Of the three studies that reported recruitment targets,^52, 78, 80^ only one met the recruitment target;^78^ the target sample size ranged from 60 to 1300 participants. Participants recruited ranged from 11 to 595 participants. One study disclosed that participants were engaged with the primary care provider or study physician to facilitate recruitment.^59^

Retention rates varied considerably across these studies. One study of adult smokers with mental health conditions reported retention of 98% at 8 weeks.^78^ The study excluded participants with poor adherence to treatment, and study compensation included USD 110 and a smartphone used. Three studies reported retention in the 70-85% range for follow-up between 4 weeks and six months.^42, 59, 80^ One study reported high lost-to-follow-up rates, with 39% of pregnant women lost to follow-up at 6-12 weeks.^52^ Four studies reported compensation or incentives for participating in the study, which was reported as ranging between USD 110 and USD 530.^41, 59, 78, 80^

### Smoking Cessation: General Population

The largest group of studies was smoking cessation interventions for the general population. ^15, 42, 43, 46–49, 51, 53, 54, 58, 61, 65, 70, 71, 74, 77, 79, 83, 85–87, 92, 97, 98 89, 91^ Of the twenty-seven smoking cessation studies targeting the general population, online recruitment methods (such as social media and online advertisements) were used exclusively in twelve studies ^15, 42, 43, 47, 48, 54, 61, 71, 74, 77, 83, 98^ and offline recruitment methods (such as flyers and clinic visits) were used exclusively in eight studies;^49, 58, 65, 70, 79, 87 89–91^ the remaining three studies used a combination of methods ^46, 53, 86^ and four did not provide information on their recruitment methods.^51, 85, 92, 97^ Six studies recruited participants through clinical settings or smoking cessation services.^49, 58, 65, 70, 78, 79^ One study, Graham et al., was a quality improvement study, meaning that participants were automatically enrolled in the study when they signed up for an online smoking cessation program. The number of participants ranged from 8 participants to 2,806 participants. Of the studies that discussed the relationship between the recruiter and participants, trained smoking cessation ambassadors (being college/university students and volunteers from non-governmental organizations) approached smokers at recruitment sites,^97^ and case managers liaised with participants in another study.^74^

Eleven studies reported a recruitment target, reflecting that the studies targeting smoking cessation in the general population were generally clinical trials.^43, 46, 47, 65, 70, 71, 77, 79, 86, 97, 98^ The recruitment target ranged from 120 to 3000 participants. The recruitment target was met in almost all studies except Affret et al. 2020,^43^ recruiting 2806 of the 3000 participants, and Rajani et al., 2021^86^ recruiting 116 of the 140 participants required.

Three studies reported high rates of loss-to-follow-up in one or both conditions.^43, 53^ In Affret et al., almost two-thirds of participants in each group were lost to follow-up in six months.^43^ In Goldenhersche et al., 80% of participants in the control group dropped out of the study after 90 days.^53^ Likewise, in Garcia-Pazo et al., 61% of the intervention group dropped out before the end of the treatment.^91^ Retention was also poor in Graham et al., where only 54% of participants completed the programme; however, the opt-out design of the study may have contributed to the high drop-out rate.^54^ Most of the other studies reported retention between 70 and 90%, with five studies reporting retention above 90% .^15, 58, 70, 74, 77^ Twelve studies reported the use of incentives provided at follow-up, which ranged from around USD 10-15 per survey to USD 150 for completing 90% of ecological momentary assessments (EMA).^41, 46, 49, 57, 58, 65, 74, 79, 86^

### Lessons Learned

Several key lessons were highlighted in the included papers related to recruitment and retention. The authors reflected on their methods and insights into recruitment and retention that are likely useful for other researchers working in mHealth interventions for addiction and problematic substance use. These are summarised in Figure 2 and described below.

**Figure 2.**
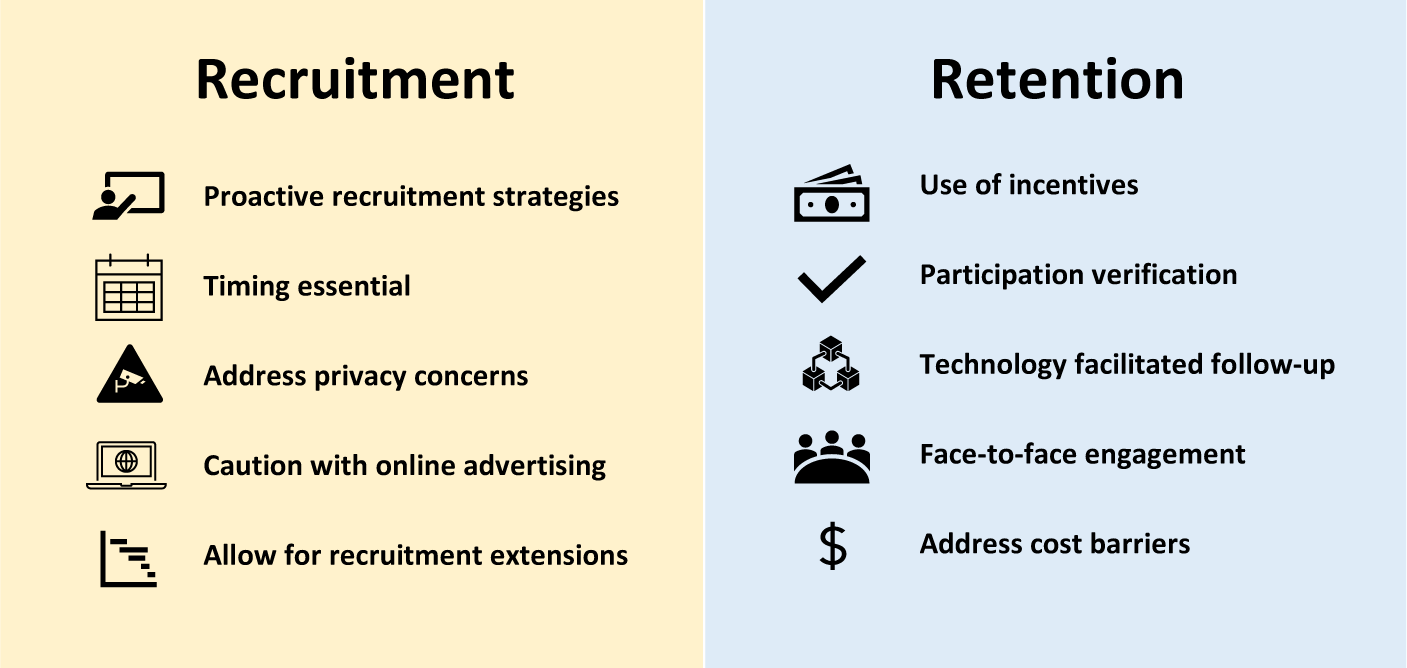
Key lessons learned for recruitment and retention.

### Recruitment

Authors noted the need for proactive and responsive recruitment strategies, such as recruitment at large public settings, including schools, ^57^ although these were not always effective. ^80^ Timing of recruitment pushes; for example, around New Year, appeared to contribute to increases participant recruitment.^55^ Provision of novel technology interventions, such as Virtual Reality (VR) and the use of a mobile Carbon Monoxide (CO) checker, also were effective in attracting participants.^53, 65^ Concerns about privacy, especially for interventions for problematic substance use, were a potential deterrent to participation. These concerns were generally found in older rather than younger participants.^95^

Internet advertisements for recruitment should be approached with caution as these methods result in large amounts of people registering interest but being subsequently disqualified.^41^ Researchers also noted the importance of knowing whether grants used to fund the research allow for the duration of recruitment to be extended, especially given the hard-to-reach nature of populations in the addictions field.^52^

### Retention

Potential facilitators of retention included participant verification, incentives, and technology. One study included safeguards to verify participant information, enabling follow-up at the intervention’s end.^41^ Incentives for completing follow-up tasks also appeared to enhance the retention of participants.^55^ Studies also noted how technology facilitated follow-ups, such as push notifications to notify participants of follow-up and the appearance of SMS on the user’s screen automatically.^15, 69^

Two studies noted that the nature of the intervention, that is, passive versus active interventions, may have contributed to poor retention and utilization ^58, 88^ Text-based interventions were discussed as active interventions with good reach and greater effectiveness compared to more passive interventions such as mobile applications which mean the user has to engage directly with the applications themselves.^68–70^ For example, tips or treatment content that are unlocked over time as an active mode of delivering the intervention content to participants.^58^

Participant engagement to increase retention was also discussed. Han et al. ^95^ highlighted that the participants with problematic substance use preferred face-to-face rather than online engagements as most were isolated from society.^95^ Furthermore, Han also stated the importance of pre-intervention education for participants before engaging with mHealth interventions. This provides the knowledge needed to engage and install a study mobile application, which could help increase their engagement with the intervention and subsequent retention.^77, 95^ Cost barriers such as mobile phone data charges are also essential to address for participants to retain them in mHealth studies.^80^

## DISCUSSION

Our review of recruitment and retention in mHealth interventions highlights the need to use various recruitment methods, especially when recruiting hard-to-reach communities. While some studies point to crucial techniques that have improved recruitment, such as partnering with community services with strong links to the target population, our analysis demonstrates that researchers should rely on more than just one method of recruitment or source when recruiting participants to mHealth interventions.

Our review also highlights the potential trade-off between recruitment and retention. In some studies, there was a high bar to participation in the study, either through safeguards (such as verified email address in Abroms et al.^41,42^) or eligibility criteria (as in Vilardga et al., ^78^) that excluded participants unlikely to engage with the intervention, which excluded high numbers of potential participants but enabled better retention of participants. In contrast, a low entry bar to participation may result in poorer retention of participants (e.g., Rodda^73^), with high loss-to-follow-up affecting the analysis and validity of findings. Several studies also showed variability in retention across groups. For example, retention rates in Goldenhersch et al. ^53^ nearly halved between Day 1 and Day 90, which the study authors attributed to high loss to follow-up in the control group (97% Day 1 vs. 57% Day 90). This highlights the importance of ensuring strong recruitment and retention techniques across all study groups.

Differential retention across control and intervention conditions suggests that alternative study designs may be needed in the digital intervention space. Clinical trial methodologies, specifically RCTs, have been explicitly designed to evaluate pharmacological interventions rather than behavioural or mHealth interventions and may not be suitable for evaluating these kinds of interventions. Other reviews have highlighted the practical issues in evaluating mHealth apps using RCTs, including limitations in keeping up with technological changes.^101^ There may also be ethical issues, such as the appropriateness of inactive controls when mHealth interventions digitize existing interventions and the risk of crossover. There is a need to identify trial designs to improve retention in mHealth interventions and adapt to evolving technology.

Technology was identified in some studies as a critical component in both recruitment and retention. Technology features such as active apps, text message messages and support for notification were seen as enhancing retention and building into existing platforms (such as WeChat) and ease of use.

### Strengths and Limitations

Our review is limited by the exclusion of unpublished and grey literature. Studies that fail to meet the recruitment targets may go unpublished, particularly where recruitment has been significantly below expected. The failure to meet recruitment targets in such studies provides valuable lessons about recruiting hard-to-reach populations without publishing negative outcomes, including lessons that there is a risk of repeating the same mistakes.

A strength of our review is the inclusion of different types of addiction and problematic substance use to compare how the recruitment and retention problem has been addressed across different domains. However, a limitation of our review was that most of the studies involved smoking cessation and primarily involved the general population with a hard to reach subgroup, which may not have generalisability to other forms of addiction and problematic substance use amongst hard-to-reach populations. In particular, we found few mHealth interventions to address problem gambling.

Our review was also limited by the amount of information provided in studies, with some describing very little information about the retention and recruitment methods used or publishing this detail in protocol papers. The low proportion of papers documenting their recruitment and retention efforts in this review demonstrates the need for journal publications to request comprehensive reporting on their recruitment and retention efforts in the methods and discussion sections of their papers if these are not published elsewhere, for example, in a published study protocol or in a clinical trial registration platform. Ensuring this documentation is available enables researchers, practitioners and the public to understand the barriers and enables recruitment and retention efforts in mHealth interventions for addiction and problematic use to learn from and develop new strategies to engage better those with addiction and problematic use patterns that we aim to help.

### Implications

Researchers need to consider the trade-off between recruiting high numbers with low retention against low numbers with high attrition when defining the eligibility and exclusion criteria for participation. For example, more stringent criteria for participants, such as specific diagnosis or adherence to treatment – can improve participant retention but limit the participants that may take part in the study, as well as limiting the ecological validity of the study as treatment adherence is likely lower outside of clinical trial settings.

Technological features that improved retention, such as text messages, should be incorporated into mHealth intervention programmes whenever feasible. However, those with limited access to such services, e.g., rural and low socioeconomic communities, may need other modalities accommodated within the mHealth space. Often, the populations with limited access to such interventions will benefit most from these interventions. Therefore, future studies should reflect on novel ways to minimize the effect of different settings and population groups to ensure that barriers are not designed into mHealth interventions.

Alternative study designs might also be required in the digital intervention space, given that the RCT was not designed for digital and/or psychological interventions and does not appear fit for purpose in these areas. As such, questions must be asked regarding designing trials to retain participants effectively. What designs can mitigate retention/recruitment issues? How do we take into account participant preference when designing trials? Perhaps there is a need to move towards stepped-wedge designs or adaptive trials, while the interventions might need greater personalization, regular feedback, and more engaging content and gamification, depending on the delivery method.

## CONCLUSION

This review has shown the importance of using various recruitment strategies for mHealth intervention studies targeting addictive disorders and substance use. Specifically, our review has shown that partnering with community services with a solid link to the target populations is a core foundation of all recruitment strategies. A failure to effectively recruit and retain study participants affects our ability as researchers to evaluate the effectiveness of new interventions. It, therefore, brings into question the validity and generalisability of study findings, which may not be a valid outcome. However, the consequences of this outcome can be far-reaching in that funding for research into the effectiveness of mHealth interventions for those living with problem addictions may become more difficult to obtain.

## Data Availability

All data produced in the present work are contained in the manuscript

## Acknowledgements

Funding: This was an investigator-initiated study funded by a grant from the Health Research Council (reference 18-237).

## Author Contributions

GH conceived of the study and developed the study design. BK, and JMcC contributed to the study design and were involved in data collection. KG and AO’S were involved in data collection. DN contributed to drafting and critical revisions of the manuscript. All authors contributed to revisions and final manuscript approval.

## Appendix 1: Search Strategy and Test Search outcome

Databases

- MEDLINE
- EMBASE
- PsychINFO
- CENTRAL
- Google Scholar (first 200 results) to check for missed publications

Terms

- gambling or gamble or alcohol* or drug* or “substance use” or addiction or ‘problem* use’ or dependenc*
- mHealth or ‘mobile health’ or smartphone or ‘mobile phone’ or ‘mobile device’ or ‘cell phone’ or ‘mobile app’ or ‘mobile application’ or ‘smartphone app’ or ‘smartphone application’ or sms or ‘text messaging’
- intervention or therapy or therapeutic or treatment or rehab*
- recruit* or attrition or retentuon Term - Google scholar:
- ∼gambling | ∼addiction | “problem use” | ∼alcohol | ∼substance
- mHealth | "mobile health" | smartphone | "mobile phone" | "mobile device" | "cell phone" | app | sms | text messaging AND intervention |
- ∼therapy
- ∼retention | ∼attrition | ∼recruitment Test Search: 01/09/21

MEDLINE

- 317 results

PsychINFO

- 312 results

= 629 total → 229 duplicates = 400 total

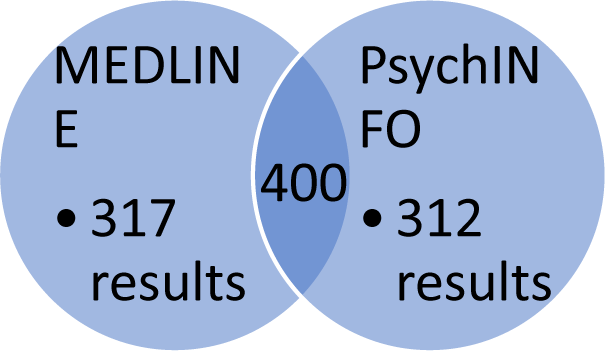

